# ACE2 polymorphisms as potential players in COVID-19 outcome

**DOI:** 10.1101/2020.05.27.20114843

**Authors:** André Salim Khayat, Paulo Pimentel de Assumpção, Bruna Claudia Meireles Khayat, Taíssa Maíra Thomaz Araújo, Jéssica Almeida Batista-Gomes, Luciana Carvalho Imbiriba, Geraldo Ishak, Paula Baraúna de Assumpção, Fabiano Cordeiro Moreira, Rommel Rodriguez Burbano, André Ribeiro-dos-Santos, Ândrea Kelly Ribeiro-dos-Santos, Ney Pereira Carneiro dos Santos, Sidney Emmanuel Batista dos Santos

## Abstract

The clinical condition COVID-19, caused by SARS-CoV-2, was declared a pandemic by the WHO in March 2020. Currently, there are more than 5 million cases worldwide, and the pandemic has increased exponentially in many countries, with different incidences and death rates among regions/ethnicities and, intriguingly, between sexes. In addition to the many factors that can influence these discrepancies, we suggest a biological aspect, the genetic variation at the viral S protein receptor in human cells, ACE2 (angiotensin I-converting enzyme 2), which may contribute to the worse clinical outcome in males and in some regions worldwide. We performed exomics analysis in native and admixed South American populations, and we also conducted in silico genomics databank investigations in populations from other continents. Interestingly, at least ten polymorphisms in coding, noncoding and regulatory sites were found that can shed light on this issue and offer a plausible biological explanation for these epidemiological differences. In conclusion, ACE2 polymorphisms should influence epidemiological discrepancies observed among ancestry and, moreover, between sexes.

## Introduction

At the end of 2019, a new outbreak caused by SARS-CoV-2 (a coronavirus) started in Hubei Province, China. The clinical condition, COVID-19, probably arose from natural selection in bat reservoirs [1]. There are currently more than five million cases worldwide, and the pandemic has been increasing exponentially in many countries since the disease was deemed a pandemic by WHO in March 2020 [2].

The lethality rate is influenced by the speed of contagion, idiosyncrasies of the affected populations according to the containment policies adopted, socioeconomic conditions and the absorption limit of the health system [3]. Due to the high rate of transmission of the virus by air and the novelty of the infection to humans, the disease has become a global emergency problem, forcing periods of social confinement to contain the pandemic, in addition to hygiene habits [4].

Epidemiological data show a tendency towards slightly greater contamination in men; however, male mortality is significantly higher [5], representing an increase from 20% to 70% in European countries, approximately 65% in some Asian countries, and, even more peculiarly, Dominican Republic citizens showed three times more deaths in men than women [6,2,7], even considering factors related to behavioral issues. Certainly, there are biological aspects that contribute to this more adverse clinical condition.

Studies indicate that cell–virus interaction is mediated by the connection of the transmembrane glycoprotein spike (S), present in the form of homotrimers on the viral surface, to angiotensin I-converting enzyme 2 (ACE2, also called hACE2), which is responsible primarily for inducing vasodilation [8].

The level and expression pattern of ACE2 in different tissues and cells can be critical to the susceptibility and symptoms resulting from SARS-CoV-2 infection [9]. Zhou and collaborators [10], using scRNA-seq datasets, classified organs vulnerable to infection as high and low risk based on their expression levels of ACE2. At the clinical level, the symptoms of COVID-19 may be related to the entry and affinity of the virus in these organs, as observed in heart failure disease and increased ACE2 expression, in which viral infection is related to a higher risk of heart attack and worse ill condition [11]. For Li and collaborators [12], ACE2 genetic variations could be crucial to the susceptibility in different cohorts and to clinical outcomes of COVID-19.

Currently, investigations of potential genetic variations that may favor or hinder interactions between the virus and the host have been conducted [13,14,15], showing a high number of codons that can, if altered, interfere with the complexity of the virus–cell interaction. It is noteworthy that the ACE2 is located on the X chromosome, causing the impossibility of heterozygosity in men. Therefore, polymorphisms in their single copy could be related to the worst outcomes observed in males [16].

Considering the above, we sought explanations for an intrinsic factor that differed between sexes and populations that may justify the differences observed in the incidence and lethality of SARS-CoV-2 infection among the different regions of the world, as well as between sexes. We analyzed global data in the 1000 Genomes Database and, in addition, we conducted studies of exomes in two population groups in the Brazilian Amazon (Indians and miscegenated), without description in public genomic banks, and we compared this information with a public databank from a population in southeastern Brazil. These comparisons are important because Brazil has a continental size and an admixed population in the North (more Amerindians among all regions), Northeast (more Africans), and South and Southeast (more Europeans) [17].

## Materials and Methods

### Analyses in the 1000 Genomes Project

The analysis was performed on data from the 1000 Genomes Phase 3 database (1000G), which comprises 84,4 million variants in 2,504 individuals from 26 different populations [18]. These populations were concentrated in five large groups: African (AFR), Ad Mixed American (AMR), East Asian (EAS), European (EUR), and South Asian (SAS).

Through the complete sequence of the X chromosome, a region between nucleotides 15620281 and 15512643 was selected in revision GRCh37.p13 (15602158 and 15494520 in GRCh38.p13) since the angiotensin I-converting enzyme 2 gene (ACE2, Gene ID: 59272, updated on 22-Mar-2020) is located on the complementary strand, covering 107639 bp. Additionally, a region of ten thousand base pairs upstream to the gene was included in the analysis so that the initial search site became 15630281 (GRCh37.p13) [18,19], aiming to search for modifications in noncanonical sites to locate minor allele frequency (MAF) and allelic differences that could be relevant. The index considered was the difference in allele frequency among all polymorphisms (SNP, INDEL and SV) contained in the study region, regardless of their global or populational frequency.

### Analyses in Amazon Natives and Admixed Population

For the allelic comparison between the populations cataloged in the 1000 Genomes database and the population not described in the respective project, we investigated a population composed of 64 Amerindians and 82 admixed individuals from the Amazon region of northern Brazil. This study was approved by the National Committee for Ethics in Research (CONEP) and the Research Ethics Committee of the UFPA Tropical Medicine Center under CAAE number 20654313.6.0000.5172. The Amerindians represent 10 different Amazonian ethnic groups that were grouped together as the Native American (NAM) group. Tribe names and geographic coordinates of the Brazilian Amazon Indian populations are presented in Table S4. The 82 admixed individuals (Brazilian Admixed Population – BAP) live in Belém city, located in northern Brazil, where, due to the colonization process, are characterized by three ancestral genetic components: European, Native American and African. This sample group is also enrolled in a broad project. Furthermore, we also compared our findings to a database of variants analyzed in a Southeast Brazilian population, the Online Archive of Brazilian Mutations (ABraOM, we represent here as ABM) [20].

### DNA Extraction and Exome Library

The DNA was extracted as described by Sambrook and collaborators [21]. The genetic material was quantified using a Nanodrop spectrophotometer (Thermo Fisher Scientific Inc., USA). The libraries were prepared using the Nextera Rapid Capture Exome (Illumina) and SureSelect Human All Exon V6 (Agilent) Kits. Sequencing reactions were run using the NextSeq 500 High-output v2 300 Cycle Kit (Illumina®, USA) on the NextSeq 500® platform (Illumina®, USA).

### Exomic Bioinformatics

The quality of the FASTQ reads was analyzed (FastQC v.0.11-http://www.bioinformatics.babraham.ac.uk/projects/fastqc/), and the samples were filtered to eliminate low-quality readings (fastx_tools v.0.13 – http://hannonlab.cshl.edu/fastx_toolkit/). The sequences were aligned with the reference genome (GRCh37) using the BWA v.0.7 tool (http://bio-bwa.sourceforge.net/). The file was indexed and sorted (SAMtools v.1.2 – http://sourceforge.net/projects/samtools/). Subsequently, the alignment was processed (duplicate PCR removal) (Picard Tools v.1.129 – http://broadinstitute.github.io/picard/), and mapping quality recalibration and local realignment (GATK v.3.2 – https://www.broadinstitute.org/gatk/ were performed. The results were processed to determine the variants (GATK v.3.2) from the reference genome. SnpEff v.4.3t, Ensembl Variant Effect Predictor (Ensembl release 99) and ClinVar (v.2018–10) were used for variant annotations.

### Databank Analysis

Information from the databank and exomes were analyzed using descriptive statistics, considering the values of allele frequencies of populations and subpopulations as the data explored comparatively. Genotypic differences between sexes in the homo/hemizygous state were calculated based on the premise that populations are in Hardy-Weinberg equilibrium.

## Results

Analyzing the polymorphisms contained in the ACE2 locus, in addition to ten thousand base pairs upstream, we found 2266 polymorphisms, of which 199 were contained in the region 5′ upstream of the gene, 85 were located in exonic regions, and the others were located in the introns.

### Polymorphisms in exonic regions that may influence disease outcome

In the exonic region, 15 SNPs of the 85 polymorphisms found in 1000G present differences greater than 1% between some of the populations (all between exons 17 to 21). Another three polymorphisms (rs889263894, rs1027571965, rs147464721) appear mainly in the Brazilian population. Nine of these exonic polymorphisms are present in the most common isomorphs (v1 and v2) and show important differences in populational frequency (Fig 1, additional data on S1 Table).

**Fig 1.**
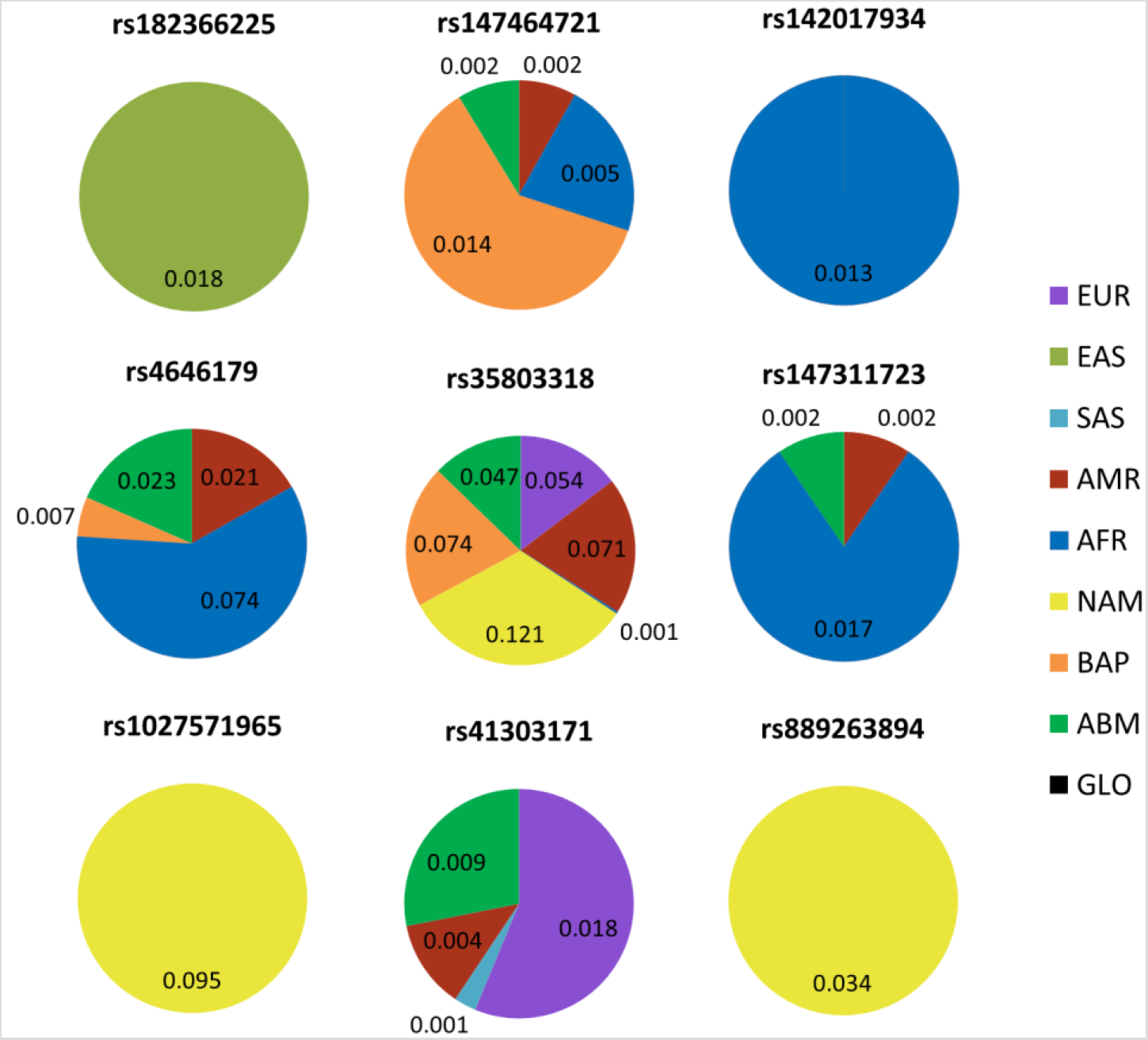
MAF of the main polymorphisms in exons present in the most common isoforms (v1 and v2) of the ACE2 gene. RS: Reference SNP; Ref: reference allele; Alt: alternative allele; EUR: European; EAS: East Asian; SAS: South Asia; AMR: Ad Mixed American; AFR: African; NAM: Native American; BAP: Brazilian Admixed Population; ABM: Online Archive of Brazilian Mutations.

As noted, some of these polymorphisms have very significant differences between MAFs from different populations. rs35803318 is absent in Asians (virtually absent in AFR) and has an average MAF of 0.05 in EUR and ABM, increased to 0.074 in BAP, and has the highest allele frequency in the indigenous population among all populations (MAF=0.121).

On the other hand, rs4646179 is absent in indigenous, miscegenated people from the Amazon, Asians and Europeans and is found in the population of southeastern Brazil and Americans, with a MAF=0.023 and an even greater frequency in Africans (MAF=0.074).

Interestingly, rs1027571965 and rs889263894 presented allele frequencies exclusively in indigenous people and are not being found in any other world population of 1000G, neither in BAP nor in the Brazilian ABM database, with MAF=0.095 and 0.034, respectively.

rs147464721 has MAF=0.014 in the miscegenated population of the Amazon (BAP); however, they did not present any allelic frequency among the indigenous population, as well as in Asians and Europeans, presenting a slightly lower MAF difference when compared to AFR and AMR.

### Polymorphisms in upstream regions that may influence disease outcome

Differences greater than 1% of MAF in the region 10,000 base pairs 5′ upstream to the gene were observed in 57 polymorphisms (Fig 2).

**Fig 2.**
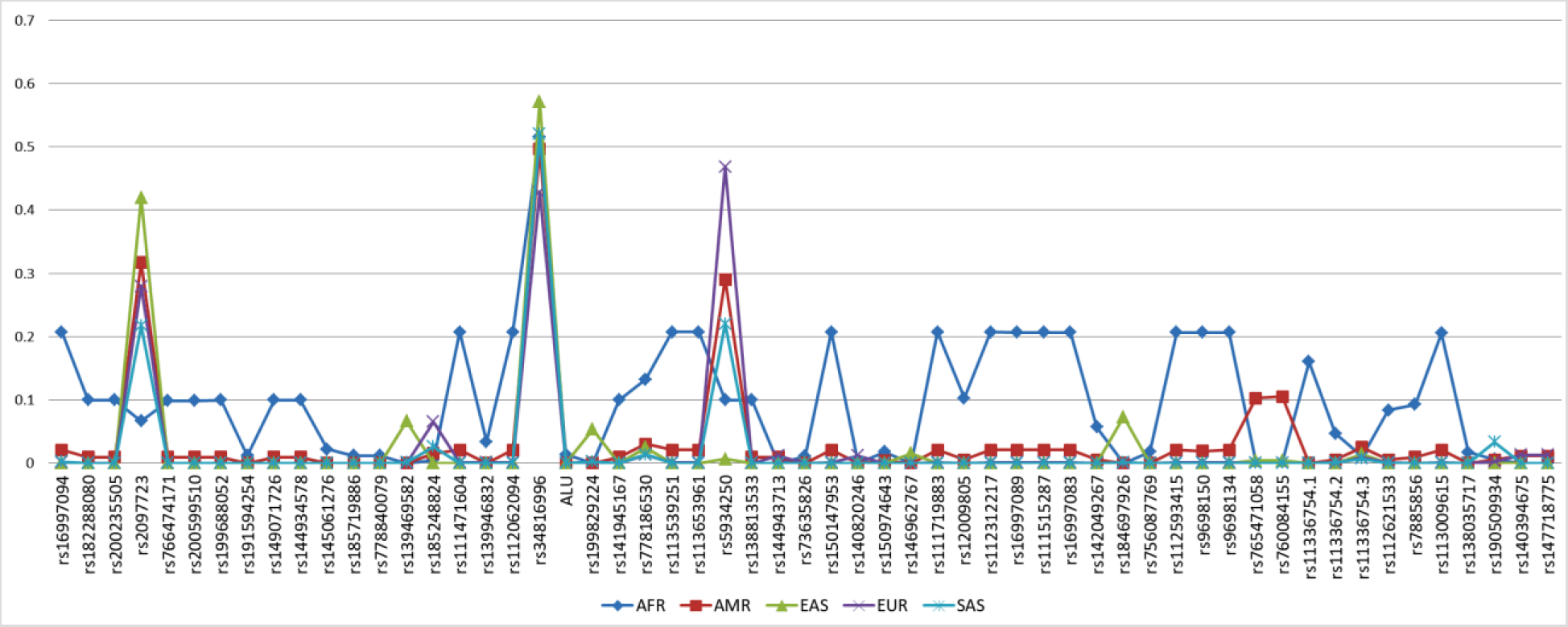
MAF of polymorphisms of the 10k 5′ upstream region of ACE 2, with MAF differences greater than 1%. Coordinate represents the frequency of the allele (MAF).

Among the highlights, rs5934250, with a change from G to T at approximately 5700 bp upstream to the gene, presented a difference of up to 0.47 in the AFR (0.10), AMR (0.29), EAS (0.01), EUR (0.47) and SAS (0.22) populations for the T allele. This means that the T allele is almost zero in the East Asian population, while it has a MAF in almost half of Europeans.

It is also worth mentioning that the rs2097723 SNP presents a very heterogeneous distribution, oscillating between 7% in Africans, 32% in Americans, 42% in East Asians, 28% in Europeans and 22% in South Asians.

### Polymorphisms in intronic regions that may influence disease outcome

In intronic polymorphisms, many of them present a very relevant MAF interpopulational difference (up to 0.46). Two of them deserve to be highlighted, rs2285666 and rs4646140, because they are near exons.

It is important to mention that rs2285666 has the highest frequency of the rarest allele (MAF = 0.71) in the indigenous population, with very important MAF differences of 0.17 (EAS), 0.23 (SAS), 0.36 (BAP), and 0.37 (AMR) and an average difference of 0.48 for the others (EUR, AFR and ABM). In contrast, rs4646140 has a MAF ranging from zero in Indians to 0.13 in Africans through EUR, AMR, BAP, EAS, ABM and SAS (Fig 3). Furthermore, considering the possibility of influence in determining isoforms v2, it is worth noting rs190614788 on intron 1 (with a difference of more than 0.11 between EUR and EAS).

**Fig 3.**
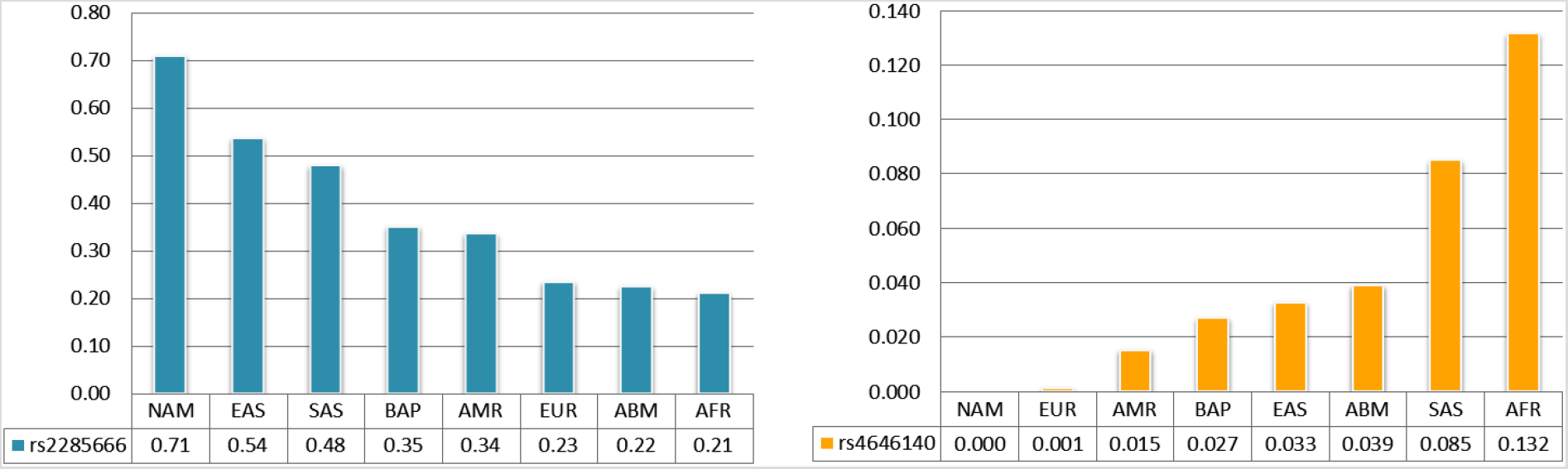
Minor allele frequency of the SNP intronic rs2285666 and rs4646140.

The main findings of our study are concatenated in Table 1 (additional information in S3 Table), and they are discussed below.

**Table 1.** ACE2 polymorphisms with the potential to influence COVID-19 and their interpopulation and inter-sex differences.

| Population* | RS | Site | MAF* | Others (MAF) | Modification (score) | Men risk | Women risk | M/W |
| --- | --- | --- | --- | --- | --- | --- | --- | --- |
| BAP | rs147464721 | codon 351 | 0.014 | absent | Synonymous | 1.4% | 0.0% | 71.4 |
| ABM, AMR, AFR | rs4646179 | codon 690 | 0.074 | absent | Synonymous | 7.4% | 0.5% | 13.5 |
| NAM | rs35803318 | codon 749 | 0.121 | EAS, SAS, AFR (null) | Synonymous | 12.1% | 1.5% | 8.3 |
| EUR | rs41303171 | codon 720 | 0.018 | absent | Asn > Asp (0.02) | 1.8% | 0.0% | 55.6 |
| AFR | rs147311723 | codon 731 | 0.017 | EAS, SAS, AMR, ABM (null) | Leu > Phe (0.941) | 1.7% | 0.0% | 58.8 |
| NAM | rs1027571965 | codon 673 | 0.095 | absent | Ala > Gly (0.045) | 9.5% | 0.9% | 10.5 |
| NAM | rs889263894 | codon 541 | 0.034 | absent | Lys > Ile (0.958) | 3.4% | 0.1% | 29.4 |
| EAS | rs182366225 | 3'UTR | 0.018 | absent | Upregulation | 1.8% | 0.0% | 55.6 |
| AFR | rs142017934 | 3'UTR | 0.013 | absent | Upregulation | 1.3% | 0.0% | 76.9 |
| NAM | rs2285666 | intron | 0.71 | EAS (0.54) to AFR (0.17) | Upregulation (Brain, Nerve) | 71.0% | 50.4% | 1.4 |
| AFR | rs4646140 | intron | 0.13 | SAS (0.085) to NAM (0) | No evidence | 13.0% | 1.7% | 7.7 |
| EAS | rs2097723 | upstream | 0.42 | AMR (0.32) to AFR (0.07) | Upregulation (Brain/Nerve) | 42.0% | 17.6% | 2.4 |
| EUR | rs5934250 | upstream | 0.47 | AMR (0.29) to EAS (0.01) | Downregulation (Brain, Nerve, Artery, Pituitary, Prostate) | 47.0% | 22.1% | 2.1 |

Population\*: Population with greater MAF; RS: Reference SNP; Site: genic region; MAF\*: minor allele frequency of population\*; Others (MAF): MAF of the other populations (or populations without MAF = null); Modification (score): Type of molecular consequence and PolyPhen Score [22]; Men risk: based on hemizygous genotypic frequency (q); Women risk: based at homozygous genotypic frequency (q<sup>2</sup>); M/W: risk ratio between sexes of being a mutant genotypically.

## Discussion

### Polymorphisms of the ACE2 gene are important in the binding region of the viral particle

Considering the regions where the virus commonly binds to ACE2 [14,15], our results point to the absence of relevant polymorphisms at these sites because many of those located in coding regions have MAFs close to or less than 0.001, as well as a very low possibility of conferring any global (or population) impact on the destination of the disease. Therefore, there does not seem to be any direct mechanism (at the sites of interaction) that could confer some form of resistance or greater propensity to contagion in humans. In view of this finding, other molecular modifications with potential functional repercussions were investigated, including 1) changes in sites distant to the viral binding locus, but which may bring about some structural protein change with the potential to influence the cell-virus interaction; 2) changes in translation regulation regions at 3′ UTR sites, at points of interaction with miRNA; and 3) modifications in transcription regulation zones in 5′ upstream regions and intragenic promoters.

### Polymorphisms with the potential to cause changes in the protein structure of ACE2 that may impact virus-cell interactions

The ACE2 gene is mainly composed of two isoforms with 18 or 19 exons (v1 and v2) that encode the same protein (805 amino acids) and three other smaller variants: x1, x2 and x3, which have rarely been studied [19,23]. Thus, we searched for genotypic information in these exons that could allow us to infer a disruption with the potential to culminate in impacts on the disease process.

Fifteen SNPs showed MAF differences greater than 1% among the studied populations, mainly belonging to exons 17, 18, 19, 20 and 21; the last two are terminal exons belonging only to rare isoforms [19].

Among these, rs41303171 is a missense SNP, causing the replacement of an asparagine (neutral amino acid) with aspartic acid (electronegative amino acid) at codon 720, which can culminate in a conformational disorder of this protein that, directly or indirectly, can change viral interactions. This polymorphic variant C is practically exclusive to Europeans (MAF=0.018), a fact corroborated by Cao and collaborators [9], mainly in British individuals (MAF=0.03). Across Asia, this allele is not found, and is only present in a single Asian resident of the UK [18]. In Brazil, ABraOM data point to MAF<0.01. Thus, the possible biological implications of this change may have some clinical-epidemiological consequences in a small niche of European patients with COVID-19 when compared to other regions of the world.

The change from leucine to phenylalanine in codon 731 (rs147311723) results in the exchange of two nonpolar amino acids that are structurally different (with the presence of an aromatic ring in this last), which may culminate in functional modifications in the ACE2 protein, with the prediction that this will occur equally to 0.941, according to the PolyPhen algorithm [22]. This polymorphism is absent in Asians and Europeans, with low frequency in Americans and Southeast Brazilians, and it has MAF=0.017 in Africans, mainly in Nigerians, with MAF=0.043 of allele A. In a study by Cao and colleagues [9], this polymorphism was described as a low frequency SNP in the 1000G database, but without any mention in the China Metabolic Analysis Project (ChinaMAP) database, reinforcing its absence in this population group.

In a context focused on the Brazilian Amazon, unprecedented data showed the presence of two SNPs absent in all populations of the 1000G. One of them is rs1027571965, which is characterized by an exchange of G>C in exon 16, leading to a substitution of alanine for glycine at codon 673 (MAF=0.095), and the other, rs889263894, an exchange of T>A in exon 13 (MAF=0.034), causing an alteration of lysine to isoleucine in codon 541; thus, this last SNP should cause structural differences by the exchange of a polar and electropositive amino acid with a hydrophobic amino acid, possibly resulting in functional changes in ACE2, with a probability of 0.958 that this event will occur [22]. Both SNPs had uncertain significance until now.

It is also noteworthy that in codons 351 (rs147464721), 690 (rs4646179) and 749 (rs35803318), there are just synonymous alterations, without any study of modifications of this enzyme in a genotype-dependent manner.

### Polymorphisms with the potential to accentuate ACE2 gene expression or translation that may impact virus-cell interactions

Our data point to a series of polymorphisms in the region upstream of the ACE2 gene, which oscillate markedly among populations and, therefore, in individuals, although some of them may not have relevant frequency at a global level. Among them, there are rs2097723 and rs5934250, with population allele differences of up to 0.35 and 0.47, respectively.

The lower frequency allele (C) of rs2097723 has a normalized effect size (NES) of up to 0.36 [24] for increased expression of the ACE gene in brain tissue. The T allele of rs5934250 has an NES of 0.64 for lower ACE2 expression in this tissue, among others. Thus, when looking at population data, it can be inferred that, according to these two variants, populations in East Asia would have the worst scenario with regard to increased gene expression and, Europeans or Africans have the most favorable allelic combination for these two SNPs in the pre-genic region.

Another interesting observation involves the exon 19 polymorphisms, which are contained in the 3′ UTR region of the two most important isoforms, the canonical miRNA binding site for translation control dependent on this epigenetic mechanism [19,25]. In this regard, observing the regions of interaction between the miRNA and nucleotide exchange sites, it can be noted that rs182366225 is included in the site of attachment to the seed region of miR-140–3p.1 and miR-483–3p.2, both with a match of 7mer-1A and Context ++ score percentile of 90 and 74, respectively. Thus, the proportion of translation regulation that depends on these miRNAs will be upregulated in the East Asian population, especially in Vietnamese and Chinese individuals, who have an average MAF of 0.032 of the C allele, which is not observed in any other population in the world [26]. A reporter assay for miR-483–3p predicted targets by Kemp and collaborators [27] showed that the regulation of ACE2 is dependent on this microRNA.

The rs142017934 site includes the connection site for miR-610 and miR-3646, both with 8mer of match in ACE2 (context ++ score percentile of 98 and 77, respectively), in addition to the seed regions of miR-3609 and miR-548ah-5p, both with a match of 7mer-m8 and scores of 84 and 74, respectively [25]. This polymorphism occurs exclusively in the population of African origin (MAF=0.013), mainly in Nigerians and individuals from Barbados with African ethnicity, with an average of MAF=0.026. This higher frequency among people of African ethnicity residing on different continents is probably due to the strong population ancestry of Barbados being from West Africa, a region containing Nigeria [28].

Among the intronic polymorphisms, rs2285666 draws much attention because it presents the highest frequency of the rarest allele (MAF=0.71) in the indigenous population, with differences ranging from 0.48 to 0.17 for the other populations, with the Asians being the most similar to the Indians, mainly the Chinese. The high MAF observed in the Chinese population in the present study corroborates the data of Cao and collaborators^9^ using the ChinaMAP database. The substitution of C for T in intron 4, to only four nucleotides of exon 3 (located in the splicing region), influences the gene expression in brain tissues and tibial nerve in some way so that the T allele is related to a significant increase in the expression of ACE2 [22], thus being a determinant in clinical differences that will be demonstrated in this naturally more vulnerable population.

Another intronic polymorphism, rs4646140, has no MAF in the indigenous population, reaching 0.13 in Africans, mainly in Nigerians (0.17). Some studies show the influence of these two intronic SNPs with hypertension [29,30], since there is a possibility that they will interfere in the ACE2 protein product.

In conclusion, considering this genetic aspect involving ACE2 in the complex relationship between SARS-CoV-2 and humans, we emphasize the following:

There are genetic markers that could influence the unequal rates of aggravation and death observed between men and women. Considering the polymorphisms as harmful when in homozygosity (genotypic frequency equal to allelic frequency squared) or hemizygosity (genotypic frequency equal to allelic frequency), men would have this conditions from 1·4 to 77 times more (uniallelic) than women (Table 1). Thus, a relevant contribution to the understanding of higher mortality in males is presented, as reported in the various populations affected by COVID-19.

The rates of contagion and death fluctuate greatly; in this sense, ACE2 polymorphisms could contribute to these differences. The rs182366225 and rs2097723 polymorphisms are more frequent in the East Asian population and are potentially unfavorable to the individual, as they increase the expression of the enzyme. These allele frequencies are even higher in Chinese and Vietnamese populations. Such markers are on the order of 30% to 180% more frequently in East Asians than in other populations.

Indigenous populations from Amazon have exclusive genetic polymorphisms (rs1027571965 and rs889263894) or with higher frequencies (rs2285666 and rs35803318) than other populations. These polymorphisms are related to increased expression of the ACE2 gene in brain tissues, among others. This is an extremely relevant finding because they may influence the outcomes in these populations, whose involvement by COVID-19 was recently reported. This population group, due to its genetic peculiarity and less previous exposure to viral infections, represents a major challenge in understanding and handling this pandemic.

Africans have higher rates of three relevant polymorphisms (rs147311723, rs142017934 and rs4646140). Polymorphism in rs142017934 is exclusive to this population and can influence the translation regulation of the ACE2 gene, thus enhancing the expression of this gene, which may present an increased risk for individuals who carry this variant. However, Europeans and some Africans have a higher frequency of an allele (rs5934250) that seems to reduce the expression of ACE2 in some tissues. Individuals with this genotype would have a more protective factor against this infection.

Therefore, the study highlights the importance of this genetic factor in facilitating or restricting infection and, especially, in potential clinical manifestations and outcomes. The investigation of these polymorphisms in patients affected by COVID-19, with different clinical conditions and outcomes, has great potential to favor understanding the behavior of this pandemic disease.

## Data Availability

All 1000G files are available from the The International Genome Sample Resource database (number 20140730, reference GRCh37.p13; https://www.internationalgenome.org).
All ABraOM files are available from the Brazilian genomic variants database (number 1.0.2, reference hg19; http://abraom.ib.usp.br/index.php).
NAM and BAP data cannot be shared publicly because of these samples group is also enrolled in a broad project, in progressing.

http://abraom.ib.usp.br/index.php

## Acknowledgments

The authors are grateful to the Federal University of Pará (Propesp), CNPq (National Council for Scientific and Technological Development) and CAPES for supporting the current research; to the 1000 Genomes Database and ABraOM for making their data available to researchers; and to all the people who, with their genetics or dedication, collaborated here to understand an important aspect of this pandemic.

## Supporting information

**S1 Table. The minor allele frequency (MAF) of SNPs in exonic regions of ACE2 that showed differences between the investigated populations**. RS: Reference SNP; Ref: reference allele; Alt: alternative allele; EUR: European; EAS: East Asian; SAS: South Asian; AMR: Ad Mixed American; AFR: African; NAM: Native American; BAP: Brazilian Admixed Population; ABM: Online Archive of Brazilian Mutations; GLO: Global (1000 Genomes). Bold exons are contained in common isoforms.

**S2 Table. Chromosome position, reference (RS) and minor allele of SNPs (MAF>0.01) in ten thousand base pairs 5′ upstream of ACE2**. *Insertion of an Alu mobile element relative to the reference

**S3 Table. ACE2 polymorphisms with the potential to influence COVID-19 and their population and sex differences**. Population*: Population with greater MAF; RS: Reference SNP; Site: genic region; MAF*: minor allele frequency of population*; Subpopulation alert (MAF): Subpopulation with greater MAF; Others (MAF): MAF of the other populations (or populations without MAF=null); Modification (score): Type of molecular consequence and PolyPhen Score^23^; Hypothetical influence: based on hypothetical biological influence in disease; Men risk: based on hemizygous genotypic frequency (q); Women risk: based on homozygous genotypic frequency (q^2^); M/W: risk ratio between sexes to be a carrier of the minor allele only.

**S4 Table. Tribe names and geographic coordinates of the Brazilian Amazon indigenous populations enrolled in this study**.

## References

01. Andersen K, Rambaut A, Lipkin W, Holmes E, Garry R. The proximal origin of SARS-CoV-2. Nature Medicine 2020; 26: 450–452.

02. WHO. World Health Organization, member, states and regions. (acessed April 2020). Avalaible from: https://who.maps.arcgis.com

03. Boccia S, Ricciardi, W, Ioannidis, J. What Other Countries Can Learn From Italy During the COVID-19 Pandemic. JAMA Inter Med 2020. (accessed Abril 2020). Available from: https://jamanetwork.com/journals/jamainternalmedicine/fullarticle/2764369 doi:10.1001/jamainternmed.2020.1447

04. Wang Y, Wang Y, Chen Y, Qin Q. Unique epidemiological and clinical features of the emerging2019 novel coronavirus pneumonia (COVID-19) implicate special control measures. J Med Virol 2020. (accessed March 2020). Available from: https://onlinelibrary.wiley.com/doi/full/10.1002/jmv.25748 doi:10.1002/jmv.25748.

05. Wenham C, Smith J, Morgan R. On behalf of the Gender and COVID-19 Working Group COVID-19: the gendered impacts of the outbreak. The Lancet 2020; 395 (10227): 846–848.

06. Chinese Center for Disease Control and Prevention. The epidemiological characteristics of an outbreak of 2019 novel coronavirus diseases (COVID-19) in China. Zhonghua Liu Xing Bing Xue Za Zhi 2020; 17;41(2):145–151.

07. GlobalHealth50/50. Global Health 50/50. (accessed April 2020). Available from: http://globalhealth5050.org/covid19/

08. Kohlstedt K, Shoghi F, Muller-Esterl W, Busse R, Muller-Esterl W, Busse R, Fleming I. CK2 phosphorylates the angiotensin-converting enzyme and regulates its retention in the endothelial cell plasma membrane. Circ. Res2002 91: 749–756.

09. Cao Y, Li L, Feng Z, et al. Comparative genetic analysis of the novel coronavirus (2019-nCoV/SARS-CoV-2) receptor ACE2 in different populations. Cell Discov 2020; 24: 6:11.

10. Zhou P, Yang X, Wang X, et al. A pneumonia outbreak associated with a new coronavirus of probable bat origin. Nature 2020; 579: 270–273.

11. Chen L, Li X, Chen M, Feng Y, Xiong C. The ACE2 expression in human heart indicates new potential mechanism of heart injury among patients infected with SARS-CoV-2. Cardiovasc Res 2020; Mar 30. pii: cvaa078.

12. Li Y, Zhou W, Yang L, You R. Physiological and pathological regulation of ACE2, the SARS-CoV-2 receptor. Pharmacol Res 2020; Apr 14:104833.

13. Othman H, Bouslama Z, Brandenburg J, et al. Interaction of the spike protein RBD from SARS-CoV-2 with ACE2: similarity with SARS-CoV, hot-spot analysis and effect of the receptor polymorphism. (accessed March 2020). Available from: https://www.biorxiv.org/content/10.1101/2020.03.04.976027v3.doi.org/10.1101/2020.03.04.976027

14. Ortega J, Serrano M, Pujol F, Rangel H.. Role of changes in SARS-coV-2 spike protein in the interaction with the human ACE2 receptor: an in silico analysis. EXCLI Journal 2020; 19: 410–417.

15. Yan R, Zhang Y, Li Y, Xia L, Guo Y, Zhou Q. Structural basis for the recognition of SARSCoV-2 by full-length human ACE2. Science 2020; 367 (6485): 1444–1448.

16. Walls A, Park Y, Tortorici M, Wall A, McGuire A, McGuire A Veesler D. Structure, Function, and Antigenicity of the SARS-CoV-2 Spike Glycoprotein. Cell 2020 pii, S0092-8674(20)30262-2.

17. Santos N, Ribeiro-Rodrigues E, Ribeiro-Rodrigues E, Ribeiro-Dos-Santos A, Pereira R, Gusmão L, Amorim Guerreiro J, Zago MA, Matte C, Hutz M, Santos S. Assessing individual interethnic admixture and population substructure using a 48-insertion-deletion (INSEL) ancestry-informative marker (AIM) painel. Hum Mutat 2010 31(2): 184–90.

18. IGSR. The International Genome Sample Resource. IGRS. (accessed April 2020). Available from: https://www.internationalgenome.org/

19. NCBI. ACE2 angiotensin I converting enzyme 2 [Homo sapiens (human)]. PUBMED. (accessed April 2020). Available from: https://www.ncbi.nlm.nih.gov/gene/59272

20. ABraOM Online Archive of Brazilian Mutations. (accessed April 2020). Available from: http://abraom.ib.usp.br/

21. Sambrook J, Fritsch E, Maniatis T. Molecular cloning: a laboratory manual. New York: Cold Spring Harbor Laboratory Press 1989.

22. Adzhubei I, Jordan D, Sunyaev S. Predicting functional effect of human missense mutations using PolyPhen-2. Curr Protoc Hum Genet 2013; 76: 7.20.1–7.20.41.

23. Chen J, Jiang Q, Xia X, Liu K, Yu Z, Tao W, Gong Han J. Individual Variation of the SARSCoV2 Receptor ACE2 Gene Expression and Regulation. Preprints 2020. (accessed March 2020). Available from: https://www.preprints.org/manuscript/202003.0191/v1

24. GTEx Portal. The Genotype-Tissue Expression (GTEx). GTEx Portal. (accessed March 2020). Available from: https://www.gtexportal.org/

25. Agarwal V, Bell G, Nam J, Bartel D. Predicting effective microRNA target sites in mammalian mRNAs. Elife 2015; 4: e05005.

26. Cunningham F, Achuthan P, Akanni W, et al. Ensembl 2019. Nucleic Acids Res 2019; 47D, D745–D751.

27. Kemp J, Unal H, Desnoyer R, Yue H, Bhatnagar A, Karnik S. Angiotensin II-regulated microRNA 483–3p directly targets multiple components of the renin-angiotensin system. J Mol Cell Cardiol 2014; 75, 25–39.

28. Gouveia M, Borda V, Leal T, et al. Origins, admixture dynamics and homogenization of the African gene pool in the Americas. Mol Biol Evol 2020; pii, msaa033.

29. Zhao Q, Gu D, Kelly T, et al. Association of Genetic Variants in the Apelin–APJ System and ACE2 With Blood Pressure Responses to Potassium Supplementation: The GenSalt Study. Am J Hypertens 2010; 23(6), 606–613.

30. Luo Y, Liu C, Guan T, et al. Association of ACE2 genetic polymorphisms with hypertension-related target organ damages in south Xinjiang. Hypertens Res 2019; 42(5), 681–689.

